# Critical Mobility, a practical criterion and early indicator for regional COVID-19 resurgence

**DOI:** 10.1101/2020.07.30.20163790

**Authors:** Marcus O. Freitag, Johannes Schmude, Carlo Siebenschuh, Gustavo Stolovitzky, Hendrik F. Hamann, Siyuan Lu

## Abstract

The sharp reduction of human mobility in March 2020, as observed by anonymized cellphone data, has played an important role in thwarting a runaway COVID-19 pandemic. As the world is reopening, the risks of new flare-ups are rising. We report a data-driven approach, grounded in strong correlation between mobility and growth in COVID-19 cases two weeks later, to establish a spatial-temporal model of “critical mobility” maps that separate relatively safe mobility levels from dangerous ones. The normalized difference between the current and critical mobility has predictive power for case trajectories during the “opening-up” phases. For instance, actual mobility has risen above critical mobility in many southern US counties by the end of May, foreshadowing the latest virus resurgence. Encouragingly, critical mobility has been rising throughout the USA, likely due to face mask-wearing and social distancing measures. However, critical mobility is still well below pre-COVID mobility levels in most of the country suggesting continued mobility-reduction is still necessary.

## Introduction

Cell phone positioning data is becoming increasingly important in the fight against COVID-19.^1^ For example, despite some ethical considerations ^2^ tracking apps are starting to emerge that inform individuals where and when they had been in close contact with an infected person so that they can self-isolate and prevent further spread of the virus^3,4^ Anonymized and aggregated cellphone-tracking based mobility data^5-7^, on the other hand, is well suited for modeling the course of a COVID-19 outbreak.^8,9^ It has been shown that mobility is a leading factor explaining the variance in the observed transmissibility across different countries and that the mobility reduction patterns are strongly correlated with decreasing new cases during the “shutdown” in the US.^10,11^. In what follows, mobility, *m* shall be defined as the county or state-level median of the maximum distance traveled by individuals during a day. It can be used to inform a spatially and temporally resolved COVID-19 analysis.

Here we report on the simple yet powerful concept of “critical mobility” *m*_*C*_ which estimates the level of human mobility *m* that a local society can engage in without triggering another COVID-19 flare-up. The derivation of critical mobility starts from measuring the association of daily new case numbers with lagged human mobility data. As both datasets are typically noisy, particularly at a local (county or below) level, a mixed-effect spatial-temporal model is employed to obtain local critical mobility maps with predictive power.

There exists a clear analogy between the critical mobility indicator and an effective reproductive number *R* of the virus equal to 1. While short-term fluctuations above the critical mobility value are acceptable, sustained mobility above the critical value will likely lead to a new outbreak in the future. Comparisons to typical pre-COVID mobility levels show where mobility needs to remain subdued until a vaccine is found. Due to the lag between human mobility and growth in cases, a simple calculation of the normalized difference of (*m* − *m*_*C*_)/(*m* + *m*_*C*_) serves as a practical indicator of future growth in cases, serving as a concrete criterion that allows for valuable time to draw up corrective measures.

## Methods

In the IBM PAIRS geospatial big-data and analytics platform,^12,13^ we have curated COVID-19 data along with a multitude of diverse data layers such as satellite images, weather history and forecasts, NO2 pollution concentrations, population density, etc., in order to facilitate scalable and integrated data exploration and model development through a uniform interface. In this work, we focus on two datasets, COVID-19 cases and human mobility data^6^, that contain intriguing relationships that we exploited to further our understanding about the spatial-temporal dynamics of this disease in the context of the United States of America at the county level. Insights gained here should be applicable worldwide and to other emerging diseases that require nonpharmaceutical intervention.

For this study, COVID-19 daily new case data, as available from Johns Hopkins University^14,15^ on the state and county level, is pre-processed by filtering outliers. We then apply an N-day rolling mean, with *N* a multiple of 7 to get rid of strong weekly signals associated with periodic weekday/weekend behavior,

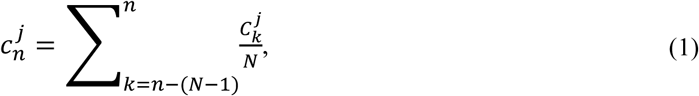

where 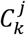 is the number of new cases on day *k* in jurisdiction *j*. The choice of *N* depends on the volatility of the COVID-19 data, and we have found *N* = 14 to yield the best results for modeling on the county level as below. (The results are almost identical when using *N* = 7 though). Figure 1(a) shows the 14-day rolling mean of new cases on the state-level. The growth rate in daily new cases is the natural log of the ratio of the smoothed new case counts 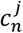 overthe previous day’s counts:

**Figure 1:**
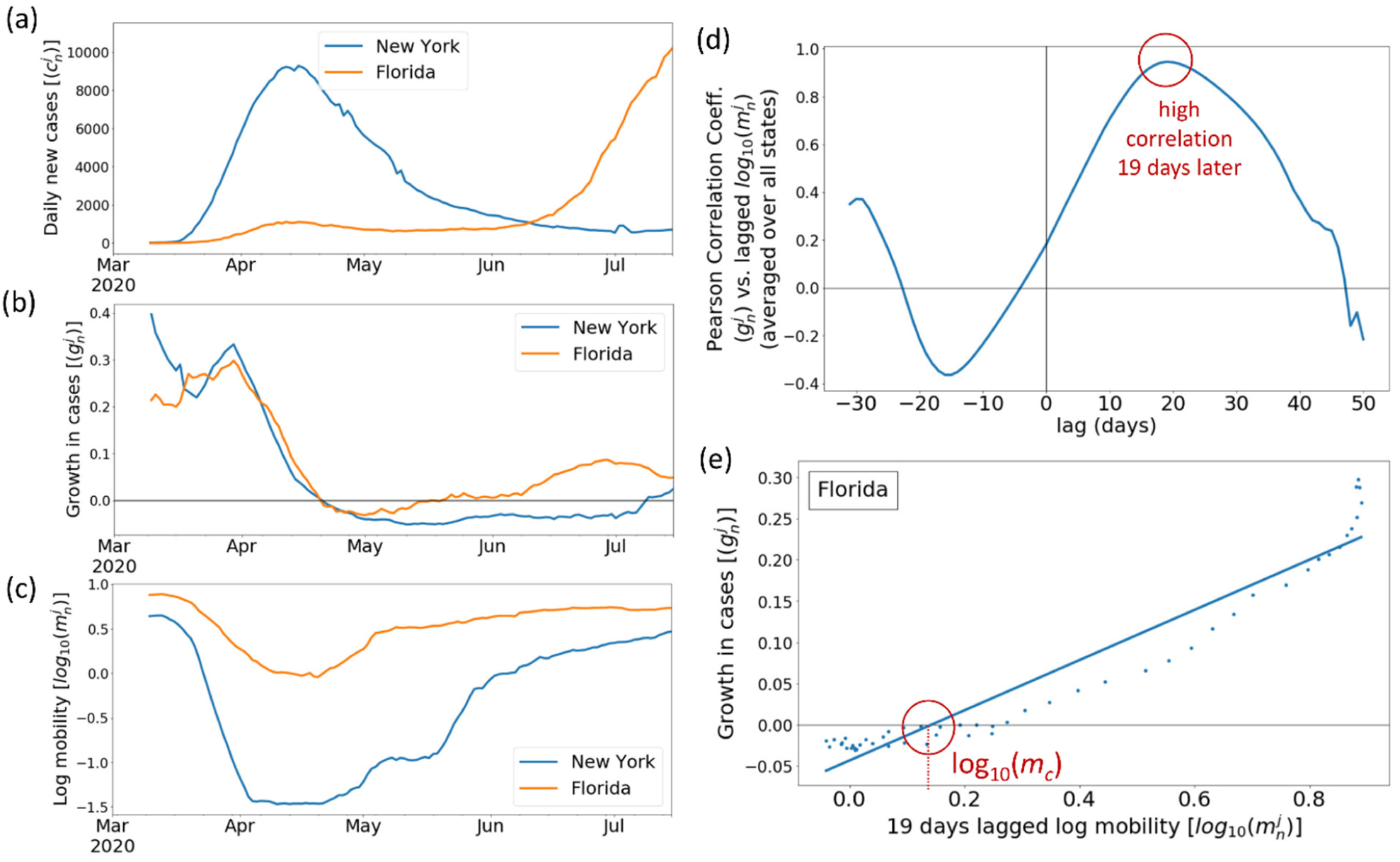
Strong correlation between (log) mobility and growth in COVID-19 cases 19 days later. (All plots are shown for 14-day rolling means as explained in the text). (a) Temporal dependence of daily new COVID-19 cases for selected US states. (b) Growth in COVID-19 cases during the same timeframe. (c) Temporal dependence of the anonymized cellphone mobility data (measured in km]). (Descartes Lab’s median of the maximum-distance mobility) during the same timeframe. (d) Lagged correlation between log-mobility and growth in cases (averaged US state-level correlations) during the abrupt shutdown phase in March/April. (e) Example of the relationship between 19 days lagged log-mobility and growth in cases during March/April for Florida with fit. The critical mobility during this shut-down phase is 10^0.14^ = 1.4 km.

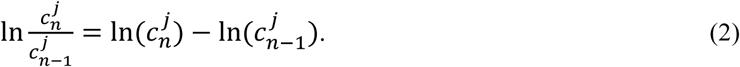

Taking the fraction above introduces significant additional noise that we smooth out by applying filtering to the daily growth data as well. (We typically use the same N=14 here as above). Therefore, the growth rate becomes

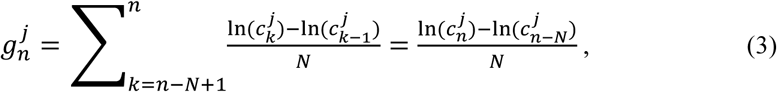

As shown in Fig. 1b for Florida and NY. The growth rate can be positive or negative and may be influenced by human mobility, social distancing, mutation of the virus, vaccinations, population immunity, and many others. Note that the growth rate can be translated into daily percentage growth *p* = 100 (*e*^*g*^− 1) or doubling time 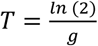.

Aggregated and anonymized mobility data from Descartes Labs ^6^ measures the median of the maximum distance (measured in km) traveled by individuals per day within a county or state by cell-phone tracking. Mobility also shows clear periodical weekday/weekend patterns and is affected by weather. However, to avoid introducing further dependencies for our model, we simply apply the N-day rolling mean for the mobility data *M* as with the above COVID-19 case data,

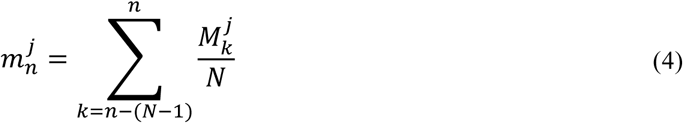

State-level data for *m* is shown in Figure 1c using *N* = 14.

We can intuitively understand that greater mobility increases the number of encounters between individuals and thus increases the risk of COVID-19 transmission from one person to another, so there should exist some dependence between mobility and COVID-19 case data. However, the lag between infection, appearance of symptoms and registration of the infection as a new COVID-19 case can be weeks and varies significantly from case to case. Given the presumably high number of unreported cases and the stochastic nature of the data it may take 2 or more steps in the chain of infections for the impact of mobility to become apparent in COVID-19 case data.

The USA shut down quite extensively at the end of March 2020. This abrupt drop in mobility imprints a recognizable change in the trajectory of COVID-19 case counts.^10,11^ Indeed, on the state-level during March and April 2020, we find a high average Pearson correlation of 0.86 (or 0.95) between log-mobility and the growth in cases 13 (or 19) days later when applying rolling windows of size *N* = 7 (or 14) days, as shown in Fig. 1d. When looking at individual state-level data we find that the lags vary by ±5 (±4) days from state to state. We are using log_10_(*m*) rather than mobility *m* here and in the following because increases of mobility by a factor are more meaningful than increases by a fixed amount when comparing different counties, for which typical mobilities may differ by an order of magnitude.

There is also a well-defined negative correlation in Fig. 1d for growth in cases about 17 days prior to the response in mobility, albeit with a magnitude less than half as strong as the positive correlation described above. So, we are in effect dealing with a feedback loop system where mobility induces future changes in growth in cases, while growth in cases induces future changes in mobility. The natural resonance periodicity associated with this feedback loop is on the order of 13+17+13+17=60 days, since only one of the two peaks is a negative correlation, and two reversals are required for a full round-trip. Damping will tend to increase this periodicity further. A similar lagged correlation plot using fatalities rather than cases (not shown) shows the anticorrelation happening near a delay of 0 days, meaning mobility responds almost instantaneously to a growth in fatalities.

From here on out, we focus on the 14-day rolling averages and the strongest peak with correlation 0.95 at a lag of 19 days as shown in Fig. 1d. We fit the relationship growth in cases () 19 days delayed vs. log_10_(*m*) on the state level, as shown for example in the state of Florida in Fig. 1e. Significant features of these fits are the slope, which reveals how much more growth in cases to expect for each additional unit of log_10_(*m*), and importantly the x-axis intercept, which corresponds to the critical point of the ln(*c*) function, where the new cases counts reach a stationary point, and the derivative (the growth in cases) is zero.

Therefore, we define a “critical mobility” *m*_*c*_ or “sustainable mobility threshold” as the mobility where subsequent growth in cases 19 days later is zero (Fig. 1e). The importance of this critical point cannot be stressed enough. Growth of zero corresponds in epidemiological terms to the effective reproductive number being *R* = 1, which means one infected person infects one other person on average. Under this scenario, caseloads stay constant over time. Mobilities that stay above the critical mobility lead to positive future growth and if not mitigated will lead to a runaway pandemic. Sustained mobility values below the critical mobility on the other hand are safer since even imported cases will eventually fizzle out at these low mobility levels. In March/April 2020 mobility in Florida fell from an initial 8 km to below 1 km (Fig. 1e). The critical mobility during this time was 1.4 km, so mobility was low enough to reduce cases at the end of April.

Table 1 summarizes all the state-level fits such as the one in Fig. 1e. The state-level slopes vary by ±50% (std) from the mean, indicating that it is difficult to assign a universal growth rate to future cases for a given mobility level. The advantage of focusing on the critical point is that it allows us to reduce the dimensionality of the problem. We are no longer asking about how much growth to expect in the future under different mobility scenarios. Instead, we are primarily interested in the mobility level at which future daily cases stay constant.

**Table 1:** Summary statistics of all the state-level fits (using 14 day rolling mean): Growth in Cases 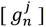 vs. 19 day lagged log mobility 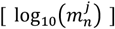 (as in Fig. 1e).

|  | count | mean | std | min | 25% | 50% | 75% | max |
| --- | --- | --- | --- | --- | --- | --- | --- | --- |
| Slope | 51 | 0.253 | 0.133 | 0.071 | 0.155 | 0.199 | 0.341 | 0.581 |
| R | 51 | 0.926 | 0.084 | 0.557 | 0.922 | 0.949 | 0.972 | 0.995 |

During the shutdown period, the mobility changes were fast and large so that mobility became almost the sole driver of changes in COVID-19 growth. During that time, it was possible to extract critical mobility values for almost every state from fits to the COVID-19 growth vs. lagged mobility data. The situation is much more complex during recovery and within finer-grained jurisdictions, such as counties. Thus, on the county level, we instead use a model informed by covariates, to smooth and fill in a sparse sampled critical mobility landscape as follows.

We regard each county’s COVID-19 and lagged mobility timeseries as an ongoing “experiment”, where the days where COVID-19 growth goes from positive to negative or from negative to positive (Fig. 2b) effectively estimates the critical mobility in that county (by sampling from the corresponding lagged mobility values) (Fig. 2a). These days can occur at a local maximum in daily COVID-19 new cases or at a local minimum. On a technical note: Due to the uncertainty of the averaged lag (estimated at ±4 days from the state-level correlations) we sample 9 different values of the critical mobility at lags from 15 to 23 instead of just one at 19 days lag and assign them to 9 successive dates. In rare cases, when there are two or more zero-crossings of the growth rate curve within 9 days, we may have multiple samples assigned to the same date (with different “lag” property). Except for the visualizations in Figs. 2c and 2d, we don’t average these but instead simply present the model training below multiple rows (samples) for the same timestamp. Sampling the data this way also reduces the sparsity of the spatial-temporal samples a bit and helps reducing day-to-day variations in modeled critical mobility.

**Figure 2:**
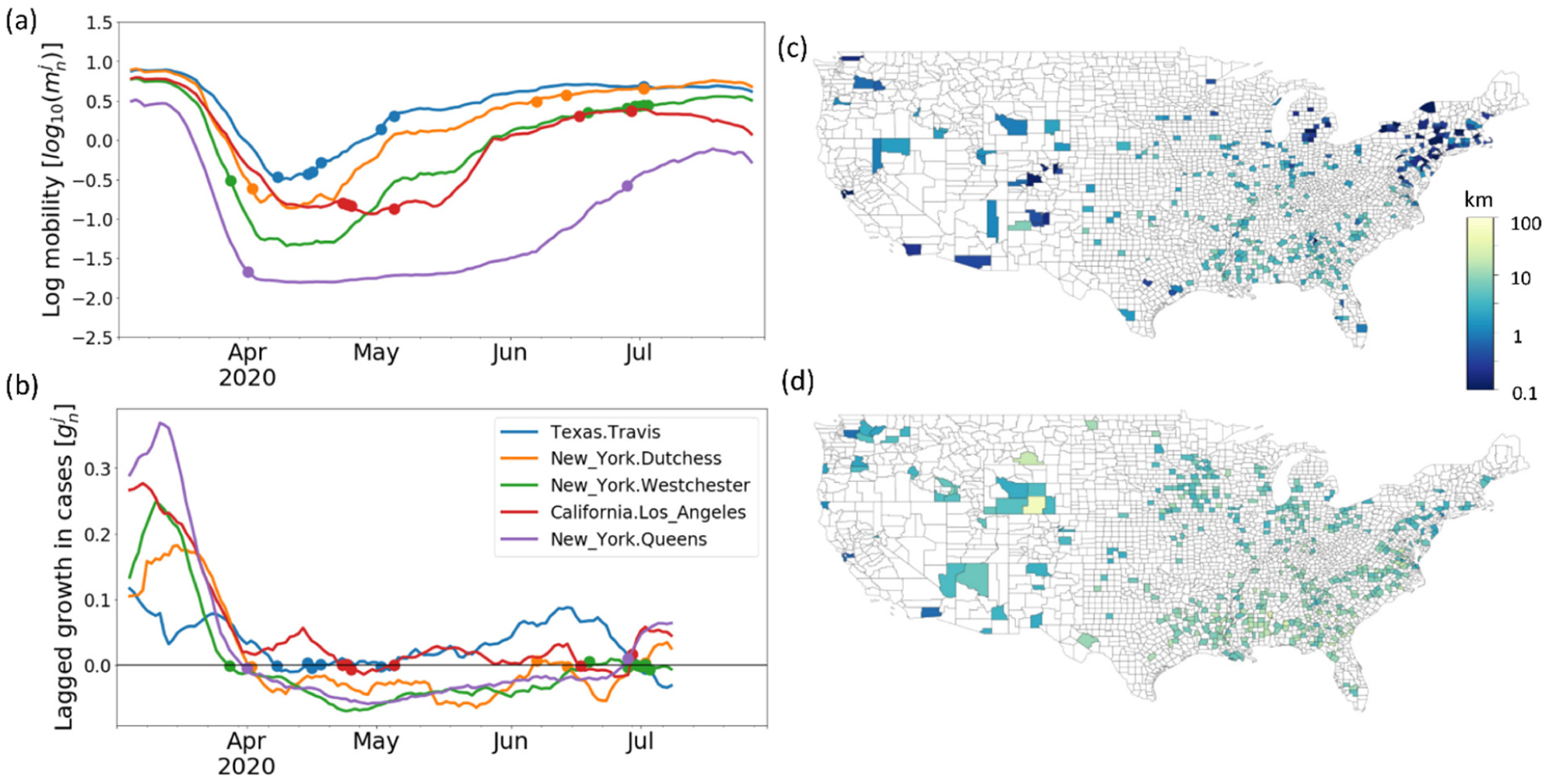
(a) Complete mobility timeseries and sampled critical mobility for a few selected counties. (b) Timeseries (shifted by -19 lag days) of the growth in COVID-19 cases for the same counties. Timestamps are sampled where growth goes from positive to negative or from negative to positive as described in the text. (c, d) Sampled critical mobility values for counties across the United States at two different timestamps. (c) April 10, 2020, and **(d)** June 07, 2020. On different days different counties’ critical mobilities are sampled. Most striking are differences between rural / urban counties as well as a general increase in critical mobility from March 26 to June 07.

Resulting spatial slices through the (sparse) measured critical mobility landscape are shown in Figures 2c and 2d for two representative timestamps, one during the shutdown and one during the re-opening phases of the COVID-19 pandemic in the USA. Below, we fit these measurements by a single spatial-temporal global model. This effectively removes outliers and helps reducing biases inherent to individually sampled data points, such as when a county is in the middle of changing testing regimes and reports biased COVID-19 growth rates. Importantly, this also allows every county’s modeled critical mobility to be informed by other counties’ experiences in similar situations.

## Results and Discussion

Several characteristics are already apparent in the measured critical mobility maps. Most obvious is the strong dependence on population density. As expected, areas in densely populated east and west-coast corridors have much lower critical mobility than rural areas in the middle of the country. In addition, rural areas close to major population centers are more impacted than rural areas far away from any major cities. This may indicate that spread of COVID-19 cases to neighboring counties plays an important role as well. Lastly, the general trend between April and June is visible throughout as an overall increase in critical mobility. It is likely that additional measures such as social distancing, face masks, avoidance of crowded indoor spaces, etc. all contribute to this positive trend in critical mobility.

In order to fill in missing areas and to smooth the volatile sampled critical mobility values, we fit a mixed model to the logarithm of the measured critical mobility data

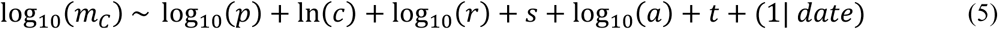

which is informed by:

1. Local population density *p*^*j*^ = *P*^*j*^/*A*^*j*^, where *P*^*j*^ is the population and *A*^*j*^ the area of county *j*
2. COVID-19 new cases (rolling mean) 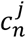 for county *j* on day *n*.
3. COVID-19 cumulative cases per capita (rolling mean): 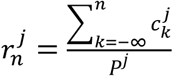 for county *j* up to day *n* (running total of daily new cases normalized by county population *P*^*j*^).
4. Non-local COVID-19 contagion 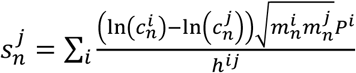 simulating the spread of transmissions from county *i* to county *j* with 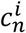 and 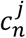 the respective COVID-19 case numbers on day *n*, 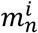 and 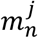 the mobilities, *P*^*i*^ the population of county *i*, and *h^ij^* the haversine distance between county centroids. The difference of the logs here is quite analogous to the difference in logs defining the growth in cases over time (eq. 2). So, in a way this term can be interpreted as the growth in cases across space due to excursions into other counties. Note that we are not modeling any drift here, such as vacationers from New York staying for an entire season in Florida. That would require much more detailed travel data. The present term assumes returning home every night.
5. A demographic feature capturing the county’s population age ratio between the 60-64 year old cohort 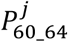 and the 20-24 cohort 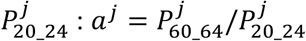
6. Temporal trend (*t*): days elapsed since January 1, 2020.
7. A temporal random effect intercept (1 | *date*), which allows capturing deviations from a linear temporal trend, caused for example by abrupt changes in human behavior that affect COVID-19 effective reproductive number R or testing availability.

Fitted critical mobility maps for two exemplary dates using this model are shown in Figures 3a and 3b. Note that we purposely keep the degrees of freedom of this model very low so that overfitting is avoided, and county-level critical mobility levels become well-informed by other counties’ experiences. This also enables the model to be easily reproduced as it only depends on three public datasets – county-level mobility^6^, daily new case numbers^14,15^, and population age distribution ^16^. We do allow for a daily random intercept (1 | *date*) to account for overall (USA-wide) changes in behavior over time, but after much experimentation, we do not allow for a county-level intercept, since our attempts to do so clearly lead to overfitting. State-level random intercepts were also problematic since they interfered with the spreading term. Parameters such as the cumulative case density in a county or the age ratio term may capture some of the county to county variability between similarly rural or similarly urban areas by for example increasing modeled critical mobility in areas with a previous outbreak that “taught” the local population how to avoid being infected through (partial) disease immunity and behavioral changes.

**Figure 3:**
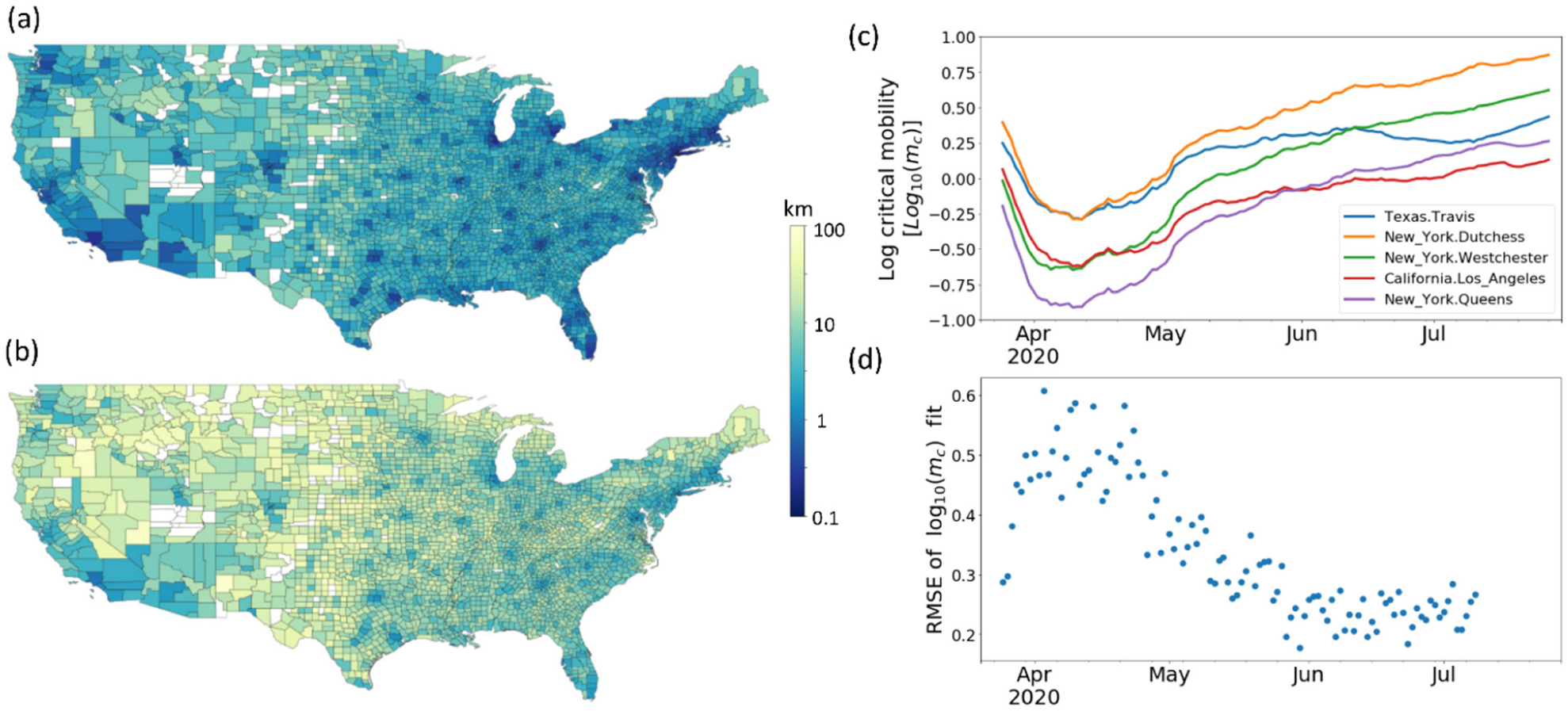
Modeled critical mobility for counties across the United States. (a) Map of mc on April 10, 2020, and (b) Map of mc on June 07, 2020. (c) Temporal dependence of log10(mc) for selected counties. (d) Grouped training RMSE (by date) showing the temporal evolution of the critical mobility model training error. One model encompassing all available counties and timestamps (with overall training RMSE of 0.35) is used in order to fit and smooth the volatile county-level data as described in the text.

Figure 3c shows the temporal behavior of the fitted (model-smoothed) critical mobility for a few selected counties. The critical mobility bottomed out in April in most counties and has generally risen since then. Throughout June and July, the heavily impacted counties in New York State have seen their critical mobility keep on rising (albeit from very low levels), while counties in the South and Southwest have not been able to increase their critical mobilities further during that time. The overall training RMSE (root mean square error) of the fit to log_10_(*m*_*c*_) is 0.35, implying that critical mobility values are associated with uncertainties around a factor of ½ to 2. As Figure 3d shows, the errors have trended down recently compared to the volatile first couple of weeks in March/April.

We consult the weights of the standardized fitting parameters in eq. (5) to gain further insights: *w*_*t*_ = 0.27, *w*_*c*_ = −0.22, *w*_*p*_ = −0.14, *w*_*s*_ = −0.05, *w*_*a*_ = 0.02, *w*_*r*_ = −0.0003. By far the most important fitting parameters are the temporal trend (*t*) (*w*_*t*_ = 0.27), number of COVID-19 cases (*c*) (*w*_*c*_ = −0.22) and the population density (*p*) (*w*_*p*_ = −0.14) within a county. Next we study each of these factors.

Apart from the first week, at the end of March when the COVID-19 outbreak in the USA was starting in earnest, the explicitly temporal mixed effect associated with t and (1|date) for the modeled critical mobility has been consistently upward (Fig. 4a). Note that this excludes indirect effects through varying case numbers. The linear trend implies an improvement of almost 2% per day in terms of critical mobility but considering the random effect’s contribution one can argue that most of the increase happened at the end of April and since then the actual trend has been up by only 1% per day. Nevertheless, it’s still a continuous improvement and we attribute it to human factors such as awareness of the COVID-19 dangers, facemask wearing habits, social distancing, avoidance of crowds especially indoors, that all reduce transmissions and increase critical mobility. The very first week is probably heavily biased due to delayed reporting of initial cases when COVID-19 testing was just getting up to speed.

**Figure 4:**
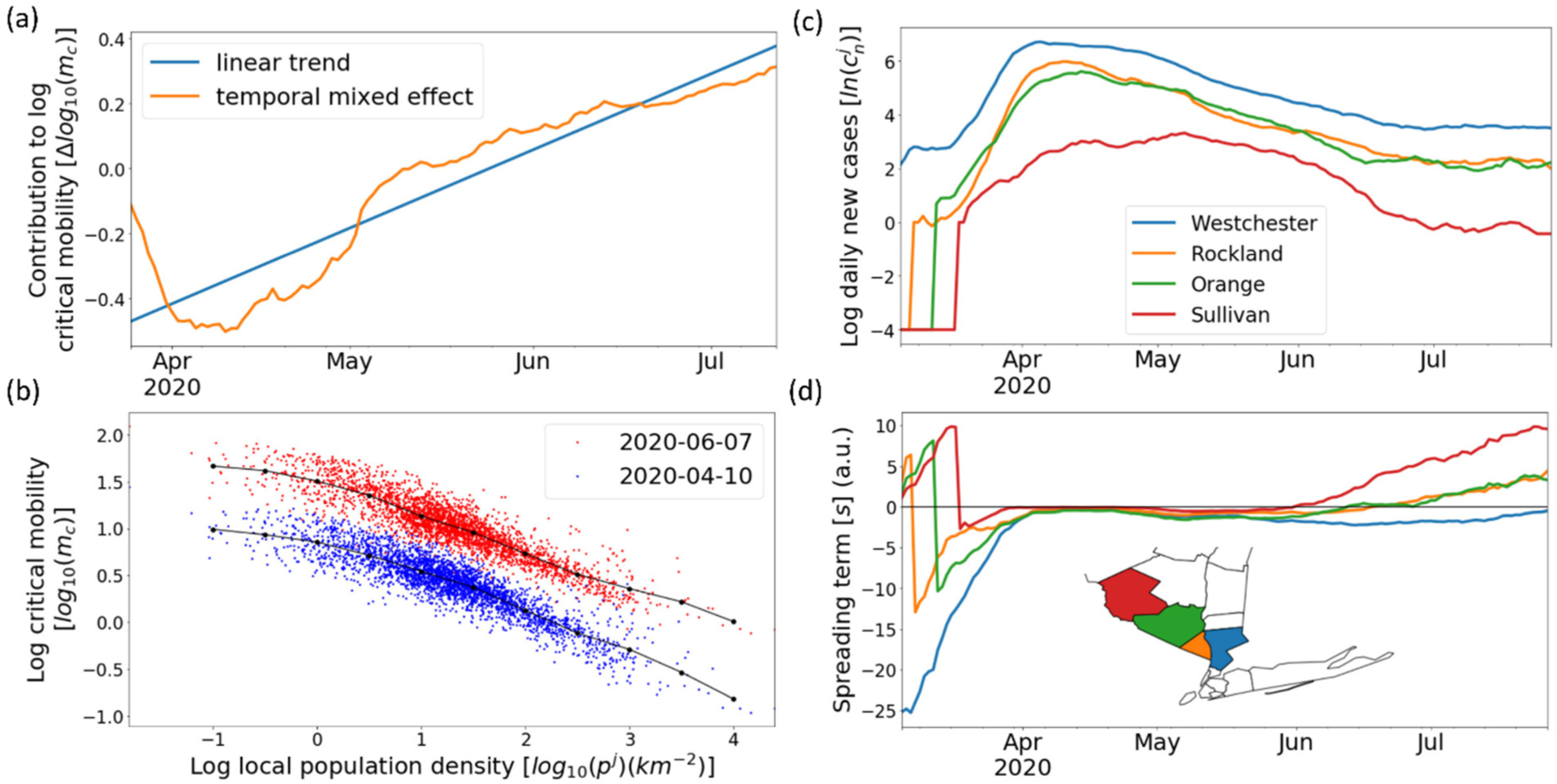
Insights from the model fitting parameters. (a) Contribution of explicitly temporal effects on critical mobility. Fixed-effect feature “trend” (blue) and trend combined with random effect “date” (orange). (b) Population density dependence of the fitted critical mobility for all counties on two dates (4/10/2020 and 6/7/2020). The black dots show the model fit using the mean of the remaining variables conditioned on the population density, time, and date. (Black lines are a guide to the eye). (c) The logarithm of daily new cases for four selected New York State counties. (d) Spreading terms for the same four selected New York State counties.

Daily COVID-19 cases have substantial contribution to the reduction in critical mobility, most likely because testing, contact-tracing, and quarantining are only effective at low enough case numbers. At medium to high COVID-19 case numbers the system for prevention gets overwhelmed, thus requiring further reduction of mobility to keep the virus in check. This is what seems to be happening since June 2020 in the southern and southwestern USA, e.g. in Fig. 3c for Los Angeles, CA, and Travis County, TX (where the city of Austin is located).

Next, we consider the population density dependence (Fig. 4b). It is intuitively clear that high population density promotes the transmission of the virus and thus decreases critical mobility, while at the other extreme, rural areas should be able to safely sustain higher mobility levels. In Fig. 4b we plot the population density dependence of the fitted critical mobility for all counties at two dates. Indeed, rural counties are more than an order of magnitude ahead of the most urban ones in terms of critical mobility. Note however that typical mobility levels need to be higher in rural areas because everything (work, shopping, etc.) is more space out.

While community spread within the same county explains most of the critical mobility data and model fit, COVID-19 spread between counties plays a small but persistent role not only during the shutdown phase (where COVID-19 cases were distributed very non-uniformly), but even during the recovery phase (where COVID-19 cases are more likely found in cities than the surrounding countryside). This can nicely be seen in the example of four New York State counties that border each other and become successively more rural (Fig. 4c and 4d). The first significant cluster of COVID-19 cases in New York State was detected in Westchester County in early March 2020. Together with New York City this formed the epicenter of the COVID-19 outbreak in New York State. It took about a week for each domino in this chain of counties to fall and begin its own COVID-19 community spread. In Eq.5 we model the spread between counties using a spreading term s that includes the weighted difference of the logs of daily new case counts between pairwise counties. Fig. 4d shows what happens prior to each of the counties starting their own outbreak: The pressure from neighboring counties in terms of the spreading term rises until the own case counts shoot up at which point the sign of the spreading term may invert and the county becomes a net exporter of transmissions. As of July, the more rural counties furthest away from New York City have seen their cases reduce significantly, but at the same time, the pressure from the urban areas nearby is rising again, limiting further critical mobility improvements. The model weight for the spreading term *s* is much lower than the weight assigned to the daily cases term *c* that is sensitive to a county’s own case count. However, in counties bordering urban areas, it may nonetheless dominate during times where the local case count is low.

The remaining fitting parameters turn out to be less important, but they might gain in importance the longer the outbreak lasts. The demographic feature (the ratio of the population in different age cohorts) 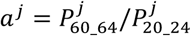 has a sign consistent with older people being more careful than younger adults. The cumulative case density feature r is intended to capture possible effects of herd immunity in counties with the highest prior infections, but as of July, it gets almost zero weight. We tried to give the model a feature of recovered population as well but that did not do any better. Some of the immunity effects may be captured by the linear USA-wide trend modeled by *t*. It will be interesting to see when local immunity effects start to play a constructive role in increasing critical mobility.

Critical mobility when compared to local maximum mobilities (which are usually observed pre-COVID), can show us quite effectively where and when local societies must remain very vigilant (Figs. 5a-b), and where they have already pushed up critical mobility enough to be on a sustainable path even without mobility reductions (Fig. 5c). Comparing Figs. 5a and 5b we can see the country-wide overall improvement, especially in the North-East, but we also see that the southern metro areas can still only afford 20% of their pre-COVID mobilities on 06/20/2020 (Fig. 5b). Note that these observations are well outside the uncertainties associated with the modeled critical mobility shown in Figure 3d.

**Figure 5:**
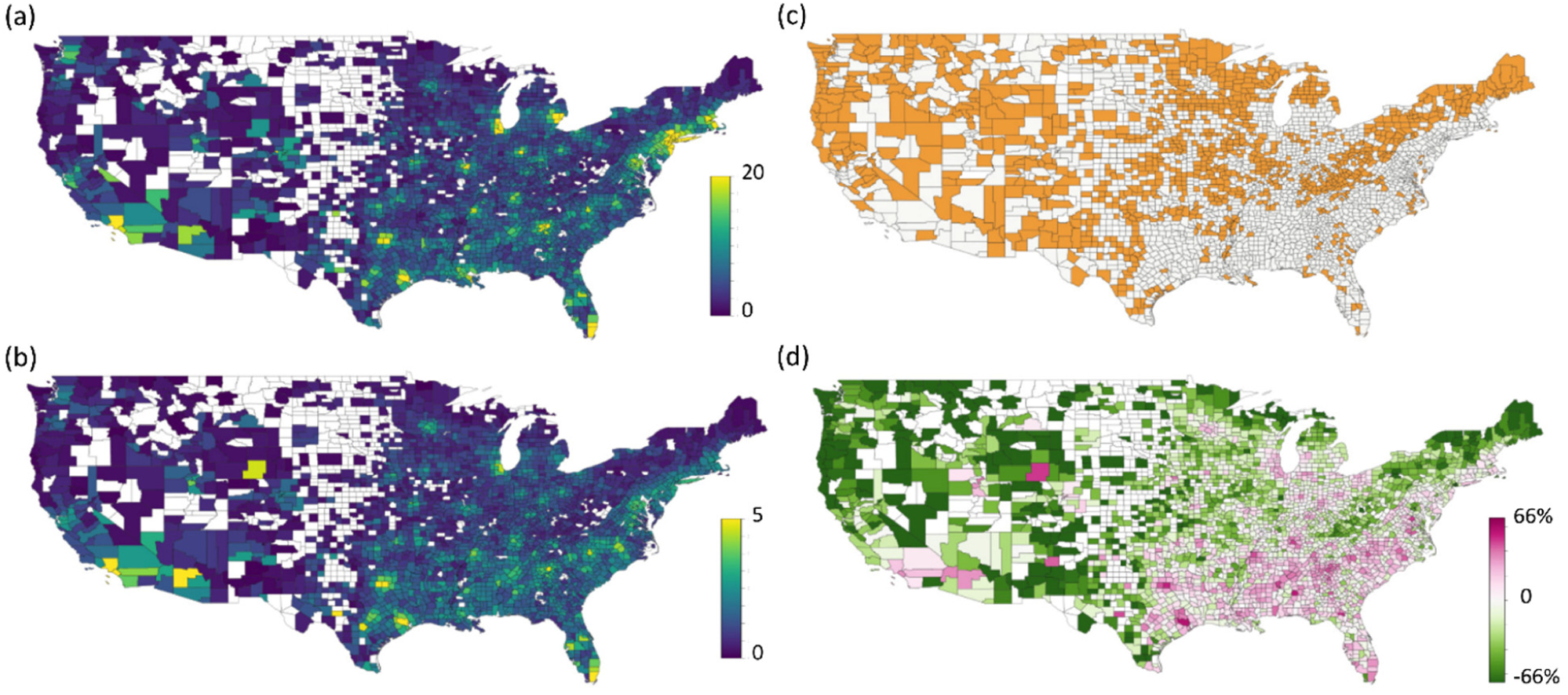
Comparison of modeled critical mobility with maximum (pre-COVID) mobility and observed mobility. a) and (b) Ratio of maximum (pre-COVID) mobility to modeled critical mobility for 2020-04-10 and 2020-06-20, respectively. (Note the different color scales.) (c) Counties highlighted in orange that have managed to push up their critical mobility (as of 6/10/2020) far enough so that their pre-COVID mobility level is sustainable. (d) Normalized Difference Mobility Index (NDMI) as defined in the text. The outlook has darkened by 6/7/2020 especially in the densely populated areas in southern California, Arizona, Texas, Florida, and the interior South.

The predictive capability of the concept of critical mobility becomes evident by comparing current mobility *m* with critical mobility *m*_*C*_. Current mobility above the critical mobility is expected to lead to future growth in COVID-19 cases while current mobility below critical mobility is expected to be sustainable. Both differencing and taking a fraction suffer when mobility values are very large or small respectively. We suggest a normalized difference mobility index (NDMI),

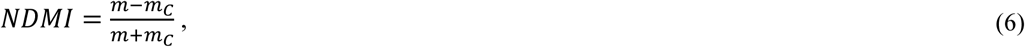

which ranges from –1 to 1, to be used as an indicator for the “danger level” a local society is engaging in, in the face of a COVID-19 situation.

Figure 5d shows the NDMI on 06/07/2020. Some of the South and South-West counties in CA, AZ, TX, and FL clearly were on an unsustainable path in early June (NMDI far greater than 0). Note that even though the critical mobility itself has been rising quite nicely during this time (see Fig. 2), the actual mobility levels have risen much faster. Northern states, that were more traumatized by the COVID-19 pandemic they experienced in April, tended to have NDMI near or less than zero.

To estimate how predictive our NDMI indicator is, we train the model until a cutoff day (May 31) and use the June 01 NDMI to infer the direction of COVID-19 growth during the following 38 days (2 times the lag time). The results are shown in Figure 6. The normalized histograms in Figure 6b show the range of outcomes one can expect for either NDMI<20% or NDMI>20%. (Note that NDMI>20% is equivalent to *m* > 1.5*m*_*c*_). The two peaks are nicely separated. Here we only considered counties where the daily new case numbers on the cutoff date are above 5 per county. For counties with lower case numbers, the prediction error increases. The accuracy of predictions using the ±20% cutoff is 0.71 and the F1 score is 0.79. For completeness, we also report an accuracy of 0.61 and F1 score of 0.69 when predicting without the ±20% cutoff.

**Figure 6:**
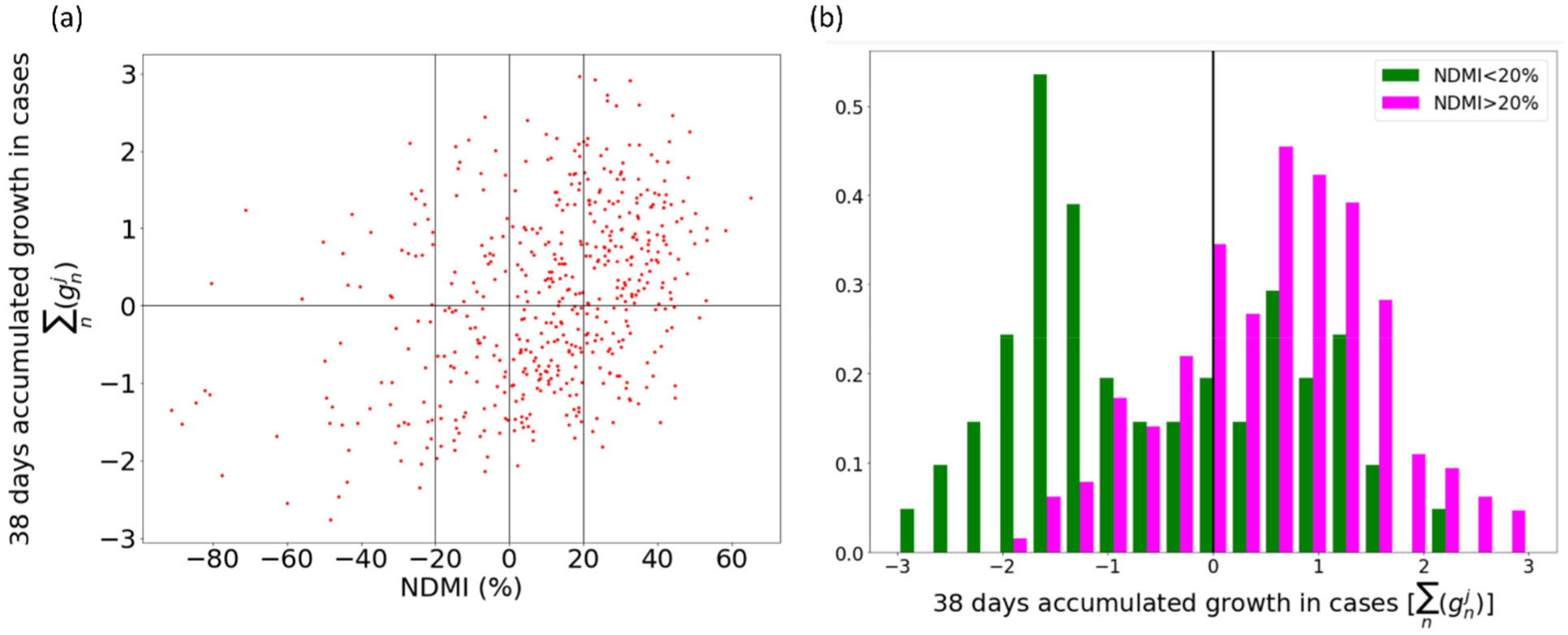
Training up to May 31, 2020, testing during subsequent 38 days. (a) Scatter plot showing the accumulated (06/01/2020 to 07/18/2020) subsequent growth in cases vs. NDMI on 06/01/2020 for counties in the USA. (b) Histograms showing the subsequent growth in cases for 2 different NDMI ranges: NDMI<20% and NDMI>20%.

While mobility was the main driver containing COVID-19 during the US shutdown^11^, it has been an open question whether mobility does still hold any predictive power on COVID-19 trajectories during the recovery time. Here we can clearly see that mobility, when compared to modeled critical mobility is still effective during June/July in identifying counties that are in danger of a COVID-19 resurgence. In addition, the critical mobility analysis can identify the subset of counties that may sustainably return to pre-COVID mobility levels.

## Summary and Conclusions

The effective reproductive number *R* of any virus largely determines its potency to spread within and between populations. For many contagious viral diseases such as the flu, *R* is initially very high, and the virus spreads rapidly until a certain percentage of the population is immune. The recovered population thus acts as the limiting factor, and once a herd immunity threshold is reached, *R* falls below 1, and the local outbreak fizzles out. In the case of the SARS-CoV-2 virus, an estimated herd-immunity threshold of around 60-70% ^17,18^ and the CDC current best estimate of Infection Fatality Rate around 0.65% ^19^ would mean 1.2 Million excess deaths in the USA alone, clearly not an acceptable outcome.

On the other hand, the 2020 COVID-19 outbreak has shown us that mobility restrictions during a shutdown are able to bring down case numbers, while the reopening of society may be associated with a renewed uptick in COVID-19 cases. In between these two regimes lies a region, where a “critical mobility” plays the role of the “herd immunity threshold” of less deadly diseases. The analogy ends there however, since the recovered fraction of the population is a monotonic function, whereas voluntary or mandatory mobility reductions may revert quickly, which leads to a renewed uptick of the effective reproductive number above 1. This balancing act results in spatial-temporal dynamics of COVID-19 flare-ups that are very different from those of the flu, which are usually well described using SIR (susceptible – infected – recovered) type of models. ^20,21^

To estimate critical mobility, we rely on a subset of counties/timestamps where growth in COVID-19 crosses zero to sample their lagged steady-state (critical) mobility. This measured critical mobility is then fitted using population density, COVID-19 cumulative case density and daily case numbers, a feature capturing the spread of the virus from county to county, a demographic feature presenting the population age distribution, a temporal trend, and temporal random effects to get daily smoothed versions of the critical mobility map that filters out the inherent volatility of individual counties’ survey data. These maps when compared to pre-COVID times show powerfully where human mobility needs to stay low or alternatively, where more needs to be done to increase critical mobility. Due to socio-economic adverse effects of restricted mobility, it is imperative that we increase critical mobility in these places by instituting additional efforts to prevent transmissions, be it through social distancing, wearing face masks, avoiding indoor spaces, or rigorous testing, contact tracing, and quarantining. Otherwise, it is likely that we will see recurring cycles of shutdowns and re-openings.

The normalized difference mobility index (*m* − *m*_*C*_)/(*m* + *m*_*C*_) reported timely within each county can be a valuable piece of information for decision making. It is a leading indicator (of at least 2 weeks) when compared to the signal from the growth in daily cases and by 4 weeks when compared to growth in daily deaths, affording us valuable time to act. An action plan could potentially be implemented using feedback loop (PID controller) theory that tries to minimize economic costs.

The model, as discussed here, depends only on three data sources: A single indicator of mobility, case numbers, and population statistics. Subsequently, it should come as no surprise that the model has a number of clear limitations. The most obvious of these might be the temporal dependence of the fit. Critical mobility essentially captures human behavior in relation to the current state of the pandemic in a given region. Thus, a model should be able to accommodate continuously changing human behavior during the course of the pandemic. However, the temporal dependence of equation 5 consists only of a linear contribution, a nation-wide random effect, and the local and remote COVID-19 numbers. As reported above, attempts to allow local changes in behavior at different times via state-wide or county-wide random effects led to overfitting. As did an independent approach to model critical mobility in a temporal rolling window. Turning from temporal to spatial limitations, the metric used when calculating spatial contributions does not take any major transport or travel routes into account, yet only spatial proximity. One can argue that this is a reasonable simplification while air travel remains limited, but this definition will have to be revisited when that is no longer the case. Finally, mobility can be defined in a number of different ways. One can imagine behavior detrimental to efforts to contain the spread of COVID-19 that will lead to very small mobility scores in terms of the metric used in this analysis.

In light of these limitations it is worth emphasizing that the key contribution of this paper is not so much the specific model, but rather the definition of critical mobility as an early indicator as well as demonstration that this indicator can be modeled in a data driven approach. Importantly, the work presented here is a model for early stages of an epidemic, where a new virus requires a non-pharmaceutical intervention, until a vaccine is available or a substantial fraction of the population becomes immune.

## Data Availability

All data used for this manuscript is in the public domain.

https://coronavirus.jhu.edu/

https://www.descarteslabs.com/mobility/

https://www.census.gov/data/tables/time-series/demo/popest/2010s-counties-detail.html

